# Is increased trunk flexion in standing up related to muscle weakness or pain avoidance in individuals with unilateral knee pain?; a simulation study

**DOI:** 10.1101/2023.12.19.23300202

**Authors:** Eline van der Kruk, Thomas Geijtenbeek

**Affiliations:** Department of Biomechanical Engineering, Faculty of Mechanical Engineering, Delft University of Technology, the Netherlands; Goatstream, the Netherlands

**Keywords:** predictive simulation, neuromuscular model, knee osteoarthritis, ageing, sit-to-walk, timed-up-and-go

## Abstract

The ‘Timed Up and Go’ test (TUG) is a widely used clinical tool for assessing gait and balance, relying primarily on timing as a measure. However, there are more observable biomechanical compensation strategies within TUG that are indicative of underlying neuromuscular issues and movement priorities. In individuals with unilateral knee osteoarthritis, an increased trunk flexion during TUG is a common phenomenon, often attributed to muscle weakness and/or pain avoidance. Unfortunately, it is difficult to differentiate between these underlying causes using experimental studies alone. This study aimed to distinguish between muscle weakness and pain avoidance as contributing factors, using predictive neuromuscular simulations of the sit-to-walk movement. Muscle weakness was simulated by reducing the maximum isometric force of the vasti muscles (ranging from 20% to 60%), while pain avoidance was integrated as a movement objective, ensuring that peak knee load did not exceed predefined thresholds (2-4 times body weight). The simulations demonstrate that a decrease in muscular capacity led to greater trunk flexion, while pain avoidance led to slower movement speeds and altered muscle recruitments, but not to greater trunk flexion. Our predictive simulations thus indicate that increased trunk flexion is more likely the result of lack of muscular reserve rather than pain avoidance. These findings align with reported differences in kinematics and muscle activations between moderate and severe knee osteoarthritis patients, emphasizing the impact of severe muscle weakness in those with advanced knee osteoarthritis. The simulations offer valuable insights into the mechanisms behind altered movement strategies, potentially guiding more targeted treatment.

## 1 Introduction

Quality of healthcare is at risk due to an increase of age-related health issues and a shortage of healthcare workers in the near future. The primary focus of many governments is to prolong the independence of older adults in their own homes. A crucial daily activity that supports this independence is the act of rising from a seated position [1]. This seemingly simple task becomes increasingly challenging as individuals age and is closely linked to a higher risk of falls [2].

In Western societies, a significant percentage of individuals over the age of 65 suffer from symptomatic osteoarthritis (OA), particularly affecting the knee joint [3]. The ‘Timed Up and Go’ test (TUG) is a clinical evaluation of gait and balance, serving as a diagnostic tool in these individuals [4]. It involves a sit-to-walk exercise where individuals rise from a chair, walk a distance of 3 meters, turn, walk back to the chair, and sit down. In a clinical setting, the time taken for this exercise is measured, if the exercise takes more than 30 seconds the patient is indicated with an high risk of falling.

Recent experimental research has demonstrated that beyond timing, biomechanical compensation strategies are indicative of the underlying age-related changes in the neuromuscular system and movement priorities in sit-to-walk [5]. Individuals with unilateral OA often exhibit asymmetrical movements while standing up, bearing additional weight on the unaffected side by leaning the trunk or by using asymmetric arm movements [6], [7], [8], [9], [10]. Additionally, they display increased maximum trunk flexion [6], [11]. Recent systematic reviews suggest that these altered trunk flexion patterns may be related to muscle weakness and/or pain avoidance [12].

Whereas experimental studies cannot distinguish between these underlying causes [13], predictive simulations are a promising tool to determine the interconnectivity and interdependency of the neuromuscular capacity, reinforcement schemes, sensory integration, and adaptation strategies in standing up. We recently published the validation of a planar neuromusculoskeletal model with reflex-based muscle control that simulates the sit-to-walk movement [14]. This communication aims to leverage this predictive neuromuscular framework to determine whether the observed alterations in trunk flexion strategies in individuals with unilateral knee osteoarthritis are more likely attributed to muscle weakness or to pain avoidance.

## 2 Methods

### 2.1 Model and controller

The design and validation of the musculoskeletal model and controller have been previously published [14]. We employed the H1120 model, featuring 11 degrees of freedom and 20 Hill-type muscles, which was initially developed as an OpenSim3 model and translated into Hyfydy (Fig. 1) [15]. Peak isometric forces in this model are based on a lower limb model by Delp et al. (1990) (G2392) [16]. The validation study showed that the simulated joint angles, muscle activation, and joint loading fell within experimental ranges with some limitations. Due to simplified contact geometries of the chair and buttocks, thighs rolling over the chair’s surface isn’t considered in the simulation which in reality extend the contact period. Ground contact forces for the stance leg’s during the initial step were underestimated due to the simplified foot contact which results in an underestimation of the ankle joint load of the stance leg during this initial step.

**Figure 1.**
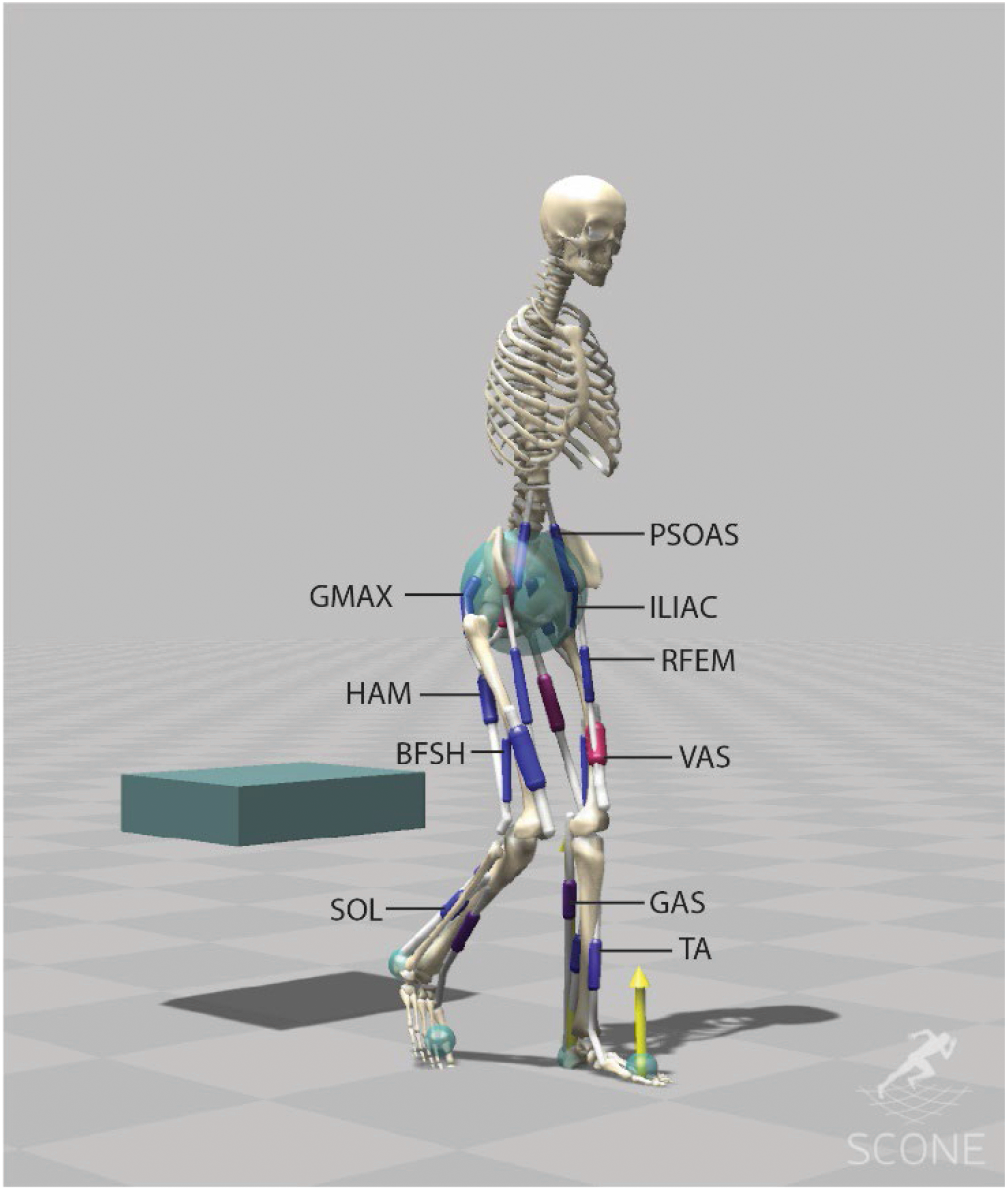
The musculoskeletal model (H1120) has 11 degrees of freedom (3 dofs between the pelvis and the ground, a pin joint (1-dof) at the hip, ankle and knee, a 1-dof lumbar joint, between the pelvis and the lumbar, and a 1-dof thoracic joint between the lumbar and the torso).) and is actuated using 20 Hill-type muscle-tendon units. The H1120 model is available on [15].

Contact forces between the feet and the ground and between the buttocks and the chair were modelled with a Hunt Crossley force spheres and box, respectively. The simulation of the sit-to-walk movement involved the utilization of a standing-up controller [14] followed by a gait controller [17]. The standing-up controller operates on a reflex control principle, featuring two distinct states, each with its own set of control parameters. The reflex controller is based on monosynaptic and antagonistic proprioceptive feedback from the muscles and vestibular feedback linked to the pelvis tilt. The timing of the transition between the states is part of the optimization problem. To account for neural latencies, we incorporated data from previous studies.

States calculated from the model were muscle length (l), muscle velocity (v), muscle force (F), and pelvis tilt orientation (θ) and velocity (θ·). The optimization problem encompassed a total of 551 free parameters, comprising controller gains (KC, KL+, KF±, Kp, and Kv), muscle length feedback offsets (lo), proportional feedback of *θ* (*θo*), proportional feedback of the lumbar and thoracic joints, transition times between the controllers, and stance load threshold for the gait state controller.

### 2.2 Objective Function

We used the objective measures from the published sit-to-walk controller [14]. These measures encompassed the following criteria: a gait velocity measure with a minimum threshold of 0.8 m/s, range penalties designed to replicate joint limitations, including lumbar extension (-50° to 0°), thorax extension (-15° to 15°), pelvis tilt (-50° to 30°), and ankle angle (-60° to 60°), a constraint on knee limit force (500 N/m) that represents the passive knee properties when the knee exceeds the range of 120° flexion or 10° extension, and minimal head acceleration (threshold of 1m/s^2). The selected speed was considered the minimum required speed to have a gait pattern that represents gait inside the lab and clinic. In daily life, normal gait speeds for older adults typically range around 0.9 m/s for women and 1.0 m/s for men [18]. As such, a speed of 0.8 m/s was considered a realistic minimum for measurements conducted in clinical environments.

Additionally, we included an energy estimate aimed at reducing energy consumption during the stand-up phase and minimizing the cost of transport during gait. The measure consisted of metabolic energy expenditure based on [19], [20] (*J*_*mb*_), and was complemented by the minimization of cubed muscle activations (*J*_*act*_), along with torque minimization ( *J*_*T*_) at the lumbar and thoracic joints to serve as a proxy for the absence of trunk muscles. *J*_*mb*_ is a sum of the muscle activation rate 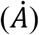, the muscle maintenance heat rate 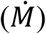, the muscle shortening heat rate 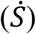, and the positive mechanical work rate 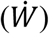. 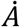 and 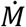 depend on the muscle mass, which was estimated by dividing the maximum isometric force over the muscle specific tension (25 N/cm^2^) multiplied the muscle density (1.0597 g/cm^3^) and optimal fiber length. Hence, when weakening the VAS muscle in the muscle weakness condition, we reduced the mass accordingly. During the optimization process, the overall energy estimate, a weighted sum between the terms (*J*_*total*_ = *ω*_*mb*_*J*_*mb*_ + *ω*_*act*_*J*_*act*_ + *ω*_*T*_*J*_*T*_) became the sole non-zero term within the objective function. We empirically set *ω*_*mb*_ = 0.01, *ω*_*act*_= 0.1 and *ω*_*T*_ = 0.0003 for all conditions.

### 2.3 Optimization algorithm

The optimization process was conducted using the open source software SCONE with the Covariance Matrix Adaptation Evolutionary Strategy (CMA-ES) [21]. Simulations were performed using the Hyfydy simulation engine [22]. The population size of each generation was 10, and the number of iterations varied depending on the simulation duration. Multiple parallel optimizations were executed with the same initial guesses based on [14], and the best set was subsequently employed as the starting point for the next set of optimizations, meaning the initial guesses for each condition were based on solution of the previous condition (-30% strength on -20% strength etc.).

### 2.4 Conditions

#### 2.4.1 Muscle weakness (VAS)

Muscular capacity progressively declines with age, with the largest relative decline in knee flexion-extension. Individuals with knee osteoarthritis also have pronounced decline in quadriceps strength [23], [24], [25]. A recent review indicates that the Rectus Femoris (RF) exhibits the most substantial relative muscle atrophy [26]. However, imaging studies and cadaver studies in older [27] and younger adults [28] show that the Vasti (VAS) muscles undergo the greatest decline in absolute PCSA. Specifically, there is an approximately 85 cm^2^ reduction in the combined vastus lateralis, medialis, and intermedius, compared to an approximate 21 cm^2^ reduction in the RF.

To therefore simulate reduced muscular capacity in knee extension, we systematically bilaterally decreased the vasti (VAS) maximum isometric force by 20%, 30%, 40%, 50%, and 60%. We chose for a bilateral approach, since the contralateral knee in subjects with unilateral OA also show quadriceps strength deficits, although in a lesser extent [29]. We initiated the model with the optimized sit-to-walk controller for the standard seat [14], [15] (neutral condition). The best set of the reduced model was then employed as the starting point for the subsequent optimization, replicating a progressive capacity decline scenario.

### 2.5 Pain avoidance

To simulate pain avoidance, we incorporated a cost function requiring the simulation to minimize the peak unilateral knee load of the stepping leg to less than 4, 3, and 2 times the body weight (BW). The joint load above the set threshold is integrated over time as measure. Individuals with symptomatic unilateral knee OA prefer to step with the affected leg first. Therefore, we implemented a cost function that becomes zero once the peak knee load on the stepping leg (left leg in simulations) was below the set threshold.

### 2.6 Measures

We analysed the trunk flexion, calculated as the angle between the global vertical and the trunk (sum of pelvis tilt, lumbar extension, and thoracic extension). Additionally, we assessed differences in timing, muscle activation, joint loading, and kinematics. The outcomes under various conditions were visualized using the OpenSim software [30].

## 3 Results

### 3.1 Reduced VAS muscle strength

Reducing the maximum isometric strength VAS led to an increased bilateral activation of the Vasti muscle (Fig. 2). The simulation compensates with increased hip extensor activity (HAM, GMAX) and increased plantarflexor muscle activity (GAS, SOL) in the stepping leg. This aligns with findings from experimental studies on sit-to-stand movements in older adults [31].

**Figure 2.**
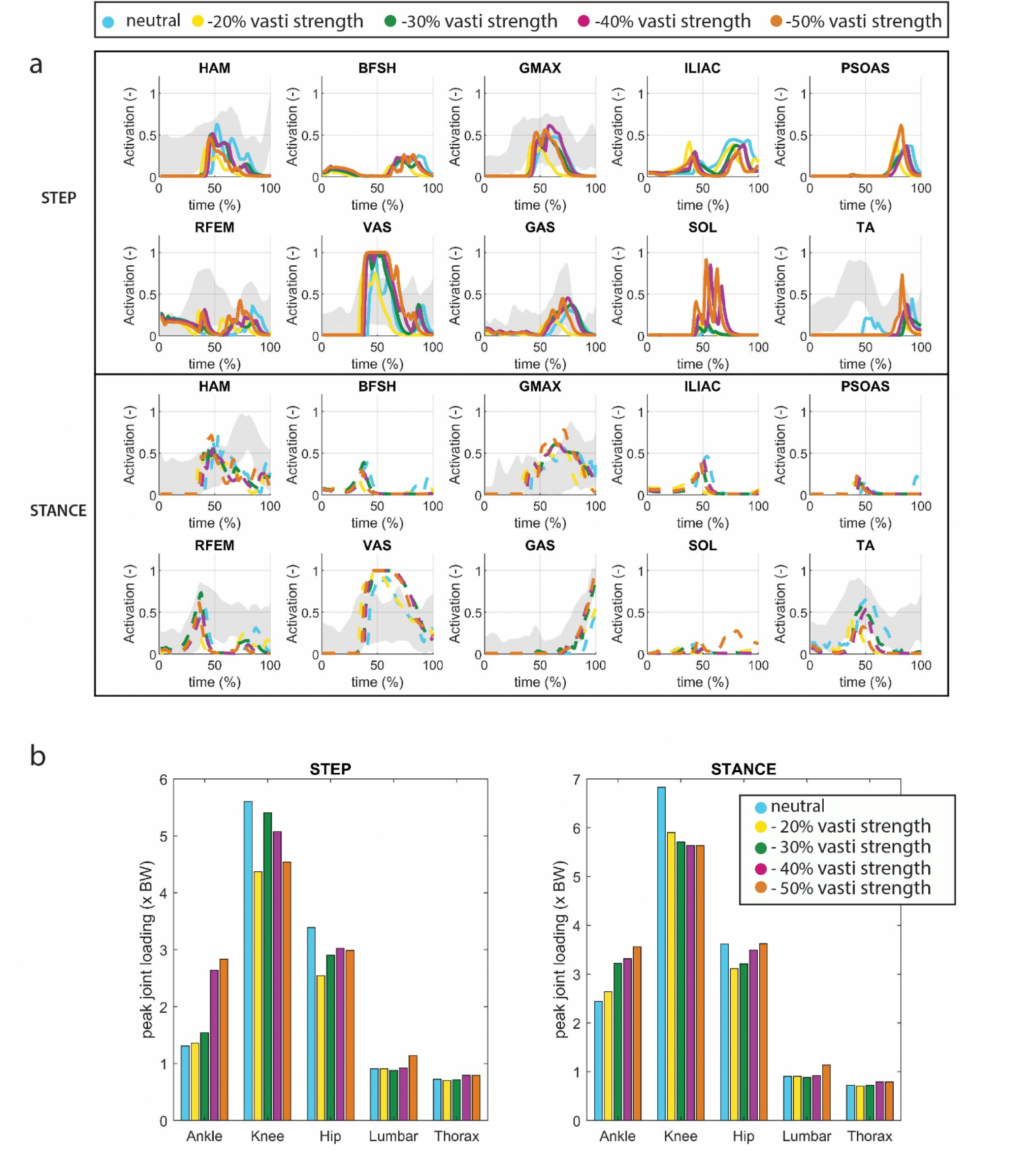
A) Muscle activation for conditions in which VAS strength is reduced. The grey shade represents experimental sEMG data adopted from [14] . Note that since no Maximum Voluntary Contraction was available in this dataset, the excitation was normalized by the maximum excitation measured within the individual complete sit-to-walk trials (stand up-walk 3 meter –turn – walk 3m – sit down). As a result the activation levels indicated by the sEMG might be higher than the actual activation levels. B) Joint loading for conditions in which VAS strength is reduced

We found a bilateral rise increase of ankle joint loading, particularly on the stepping side which doubled in the -50% condition compared to the neutral condition. Reduced VAS muscle strength thus resulted in elevated ankle loads. When VAS strength was further reduced to -60%, the model could not find a solution to rise from the seat unaided.

A minor increase in trunk flexion was observed when VAS strength was reduced (Fig. 3). Trunk flexion at seat-off reached 42.8 degrees in the -50% condition versus 38.3 degrees in the -20% condition (40.8 degrees in the neutral condition). The timing of the movement (Fig 3c) showed no noticeable pattern with reduced muscle strength. We hypothesize that in the -20% and 30% condition, there remains sufficient physiological reserve to allow for variations in gait velocity. Consequently, standing up can be slower, as gait velocity can compensate to achieve the desired average speed. Evidently, this compensatory strategy appears to be more efficient in terms of our cost assessment. However, as the strength reduction is further decreased to 40% and 50%, patterns of reduced movement velocity begin to emerge.

**Figure 3.**
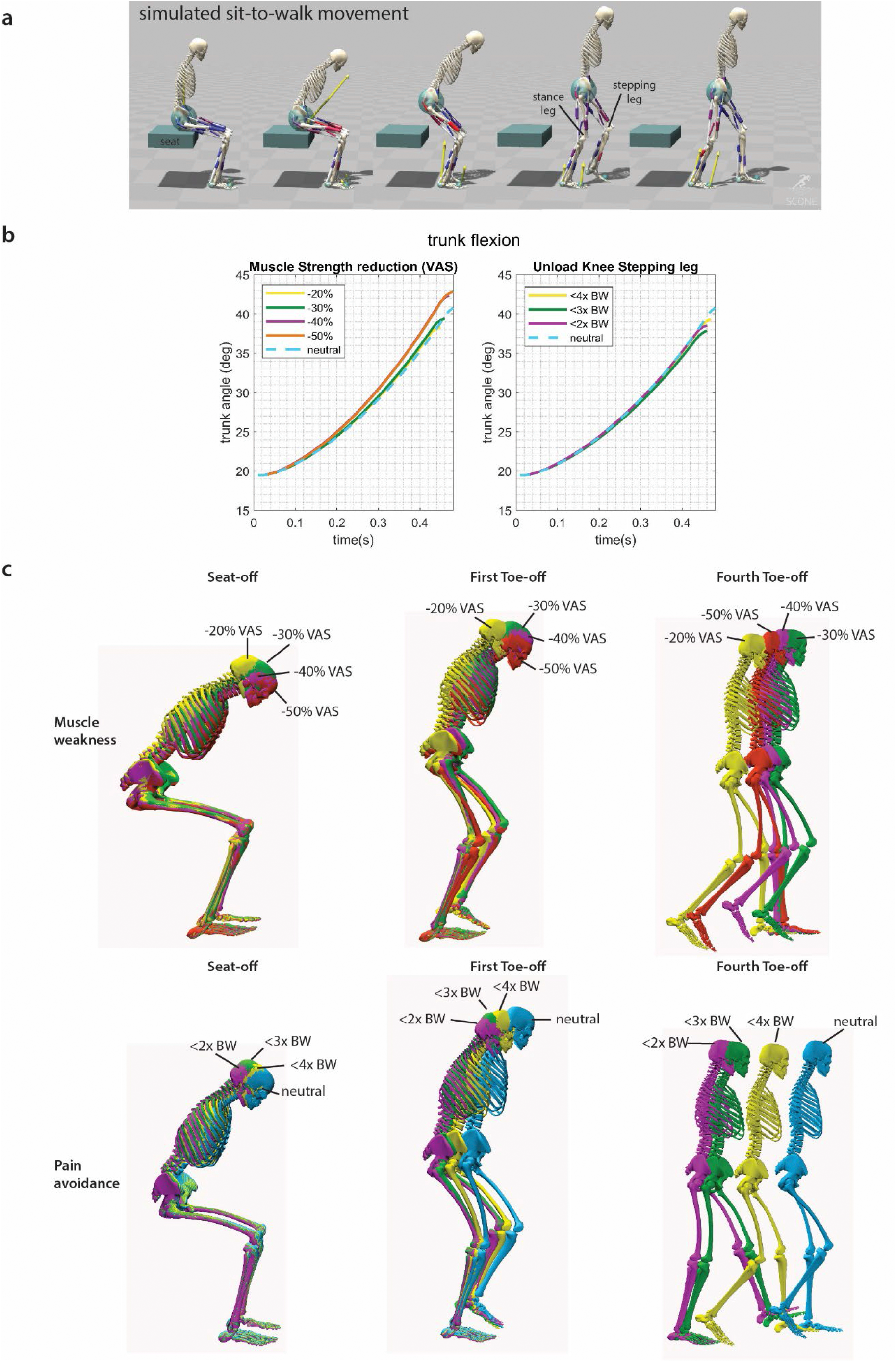
a) Simulated sit-to-walk movement adopted from [14]. The stepping leg is the leg that steps first. b) Trunk flexion from the initiation of the movement to initiation of seat unloading . c) Conditions for the sit-to-walk simulations shown in color for seat-off, first toe off (when the ground reaction force of the stepping leg becomes zero), and second toe off (when the ground reaction force of the stance leg becomes zero). There is a mild increase in trunk flexion when the VAS strength was reduced. When the knee is unloaded there is a slower movement speed.

### 3.2 Pain Avoidance

In the neutral condition, the peak knee loads (stance leg: 6.8 BW, stepping leg: 5.6 BW) and hip loads (stance leg: 3.6 BW, stepping leg: 3.4 BW) manifest just after seat-off. The peak ankle load in the stepping leg occurs during the rising phase (stepping leg: 1.3 BW), and for stance leg, it arises at the end of the single stance phase (stance leg: 2.4 BW), consistent with experimental data [14]. Unloading of the knee is accomplished by reducing ipsilateral activation of VAS and hamstrings (HAM) (Fig. 4). Following Toe-off, there is an increased ipsilateral activation of the tibialis anterior muscle (TA), resulting in increased dorsiflexion, which compensates for reduced foot-ground clearance. Unloading the knee (ranging from -29% to -59%) led to reduced load on the ipsilateral hip (ranging from -23% to -39%) and ankle (ranging from -37% to -40%). However, the ipsilateral ankle did not exhibit proportional unloading, unlike the other joints. Loading on the contralateral knee and hip increased, ranging from +5% to +11% and 0% to +5%, respectively. As for the contralateral ankle, there was a load decrease in the <4BW condition (-4%) and an increase in the other conditions (14% and 29%).

**Figure 4.**
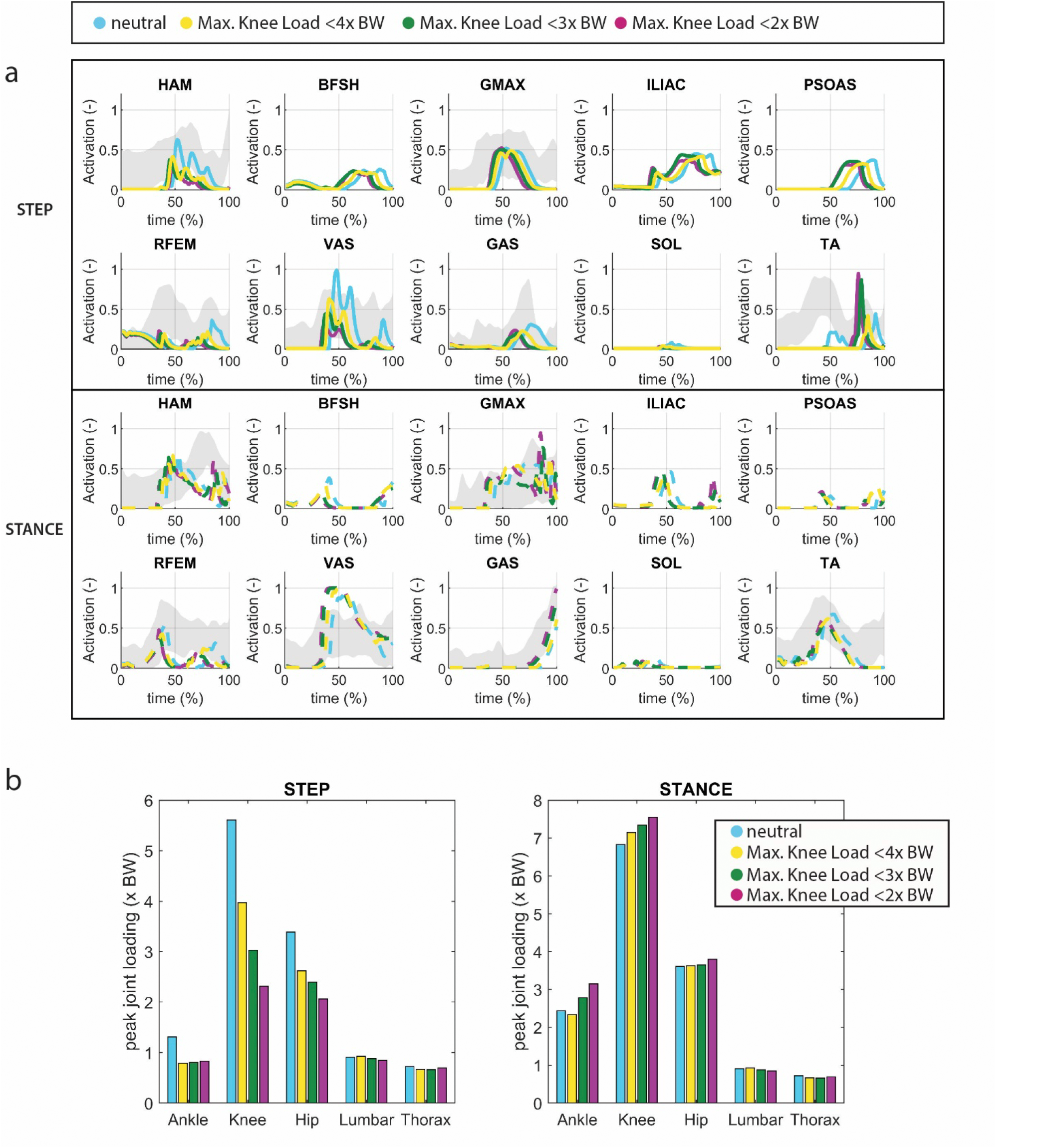
A) Muscle activation in which the knee joint of the stepping leg is unloaded (pain avoidance). The grey shade represents experimental sEMG data adopted from [14] . Note that since no Maximum Voluntary Contraction was available in this dataset, the excitation was normalized by the maximum excitation measured within the individual complete sit-to-walk trials (stand up-walk 3 meter –turn – walk 3m – sit down). As a result the activation levels indicated by the sEMG might be higher than the actual activation levels. B) joint loading in which the knee joint of the stepping leg is unloaded.

Interestingly, during knee unloading, the simulation yielded a smaller, rather than a greater, trunk angle at seat-off compared to the neutral condition (Fig 4). There was an increase in movement time observed with increased knee unloading (from initiation to heel strike of the first step: 1.1s in the neutral condition up to 1.3s in the <2x BW knee load condition) (Fig 3d).

## 4. Discussion

Our simulations reveal a minor increase in trunk flexion (2^v^) in response to a decrease in maximum isometric force in VAS (representing reduced muscular capacity). The data files, simulations, and videos can be found in the repository [32] . Experimental studies have reported an approximate increase of 9 º in trunk angle during sit-to-stand movements in individuals with osteoarthritis (OA) compared to controls (46.4 º versus 37.5º) [6]. We observed similar differences in trunk angle when simulating a low seat condition presented in [14], [15] (51 º trunk angle, approximately 10º difference with neutral condition) .Low seat height increases the task demand resulting in less muscular reserve. In essence, it reflects a reduced muscular capacity[13]. Consequently, we suspect that the trunk angle would have exhibited greater flexion had the muscle strength of more muscles been compromised.. Pain avoidance, simulated with the objective of reducing knee load, resulted in altered muscle recruitments, characterized by reduced ipsilateral activation of VAS and HAM, and slower movement speeds. However, it did not yield greater trunk flexion strategies. The predictive simulations thus support the idea that increased trunk flexion is more likely a consequence of insufficient muscular reserve than a pain avoidance mechanism.

The increase in time to perform the sit-to-walk movement is consistent with findings in the sit-to-stand (STS) task. Previous studies have reported that individuals with knee OA take significantly more time to complete the STS task and TUG compared to older adults [6], [33], [34] . Various authors have postulated that this time difference may be attributed to increased forward body bending [33]or quadriceps muscle weakness [35]. However, the simulations show that reducing movement velocity alone results in decreased peak knee loading, and might therefore be the result of a pain reduction strategy.

Our simulation model contains a number of limitations, which can be addressed in future studies:

- The simulations are conducted with a planar model. In reality, human movements during sit-to-walk involve lateral trunk motions and arm support [5]. Individuals with unilateral symptomatic knee osteoarthritis can use this to redistribute the load [12]. However, this aspect could not be tested in the current model. Additionally, individuals with knee OA exhibit increased antagonist muscle activation [8], [34], [36], [37], which is linked to the medial-lateral and internal-external rotational stability of the knee. These complexities were not captured by the simplified planar joint in the model.
- In the simulations we only tested the effect of reduced VAS isometric strength. In reality, OA patients experience muscle weakness in multiple muscle groups, which could contribute to greater trunk flexion. Furthermore, regarding the musculoskeletal model, the maximum isometric forces are based on [38], where VAS muscles are relatively weak for a healthy adult, as indicated by the maximum activation of this muscle during chair rising [14].
- The H1120 model represents a male adult with a height of 1.80m and a mass of 75 kg. Body height and mass influence movement strategies in sit-to-walk. [14]To ensure direct comparisons with experimental datasets, it would be beneficial to diversify musculoskeletal models and simulations.
- The feet were modelled using single rigid bodies, and the addition of a separate toe segment might allow for further unloading of the knee. We do not expect this to drastically change the results, because the placement and stiffness of the contact spheres allows for mimicking toe rolls.

While the Timed-Up-and-Go test is a commonly employed clinical assessment, only the timing of the exercise is typically reported in clinical and literature contexts. There is a scarcity of biomechanical data pertaining to OA patients’ performance in this test, particularly regarding which leg they use to step out first. Our choice to reduce the peak load in the stepping leg was informed by clinical observations. Given that in the sit-to-walk movement, individuals must unload one leg to initiate gait, this assumption appears valid from a motor control perspective. We did conduct simulations where the stance leg was unloaded (available online [32]). Those simulations demonstrate that the energy objective costs were higher when the stance leg was unloaded compared to the stepping leg, providing further support for the idea that unloading the stepping leg is a more natural movement.

When examining published experimental data, greater trunk flexion was only observed in individuals with severe knee OA [12]. In contrast, individuals with moderate knee OA exhibited a similar movement trajectory to that of healthy individuals but with altered muscle recruitment patterns [12]. This discrepancy can be attributed to variations in muscle strength. It is plausible that individuals with severe OA, due to reduced daily activity, possess less muscular reserve compared to individuals with moderate OA. Consequently, severe OA patients may resort to increased trunk flexion during the process of standing up as a compensatory mechanism to counteract their strength limitations.

Taking this line of reasoning further, it’s conceivable that in individuals with early and moderate symptomatic knee OA, unloading the knee to alleviate pain could potentially contribute to the development of muscular weakness in specific muscle groups, such as the vasti, over time. Humans stand up over 60 times a day and the simulations indicate that unloading the knee during this action is accomplished through reduced activation of the VAS. Since it is reasonable to infer that this compensation mechanism occurs in various daily activities, we can postulate the following hypothesis: individuals with early and moderate OA may unload the knee to mitigate pain, inadvertently leading to the underuse of specific muscle groups, particularly the vasti muscles. Over time, this underuse could result in a decline in vasti strength, resulting in increased trunk flexion, as observed in individuals with severe knee OA. Our interpretation for unilateral OA patients is that reduced activity leads to the bilateral muscle strength deficits while the specific compensation strategies during these activities, like reduced VAS activation, result in the unilateral muscle strength deficit in specific muscle groups in the affected limb (Fig 4).This underscores the significance of monitoring and addressing pain in individuals with early and moderate OA.

## Data Availability

All data produced in the present study are available upon reasonable request to the authors

https://github.com/BODIES-Lab-TU-Delft/STW-case-study-unilateral-pain.git

## Author Contributions

Eline van der Kruk – Conceptualization, Data Curation, Methodology, Formal Analysis, Funding, Validation, Acquisition, Supervision, Writing – Original Draft Preparation

Thomas Geijtenbeek - Software, Writing – Review & Editing

## Data Availability Statement

The source code and data for the predictive framework and the results presented in this manuscript are available from Git repositories: [15], [32]

